# Peer Facilitation as a Methodological Condition: What Participatory Photovoice Revealed About Transition to Adulthood Among Youth Living with HIV in India

**DOI:** 10.64898/2026.09.02.26361557

**Authors:** Anusha Sulladmath, Siddha Sannigrahi, Suhas Reddy, Meghana Gowda, Michael Babu Raj, Satish Kumar SK, Lakshmi Ganapathi, Anita Shet

## Abstract

Transition readiness instruments for youth living with HIV were developed in high-income settings and define readiness as clinical competence and individual autonomy. Few reflect youth in low- and middle-income settings, where family obligation and community stigma shape the passage to adulthood. In March 2026, sixteen youth living with HIV aged 13-22 years took part in a peer-facilitated photovoice study at two sites in Karnataka, India: Bengaluru and Belgaum. Participants photographed and collectively interpreted their experiences of growing up and their expectations of the future. Six returned in May 2026 for a reflective discussion of the method. Participants defined independence as the capacity to sustain others, located the principal risks of transition in relational and social rather than clinical domains, and recommended that transition preparation begin around age twelve. Reciprocal obligation was absent from the initial codebook and emerged through participants’ visual metaphors. Peer facilitation was a condition of the inquiry rather than an enhancement of it, and photovoice reporting should specify who exercised interpretive authority. Participatory photovoice can broaden prevailing definitions of transition readiness and inform more youth-responsive assessments.

## INTRODUCTION

The transition from pediatric to adult healthcare for youth living with HIV (YLHIV) requires not only a change of clinic but a reorganization of support systems, disclosure decisions, medication self-management, at a time of significant developmental change.^1^

India has an estimated 72,000 children and adolescents living with HIV, a population that remains underexamined.^2,3^ Pediatric care is delivered largely through vertically financed programs that concentrate psychosocial support and caregiver engagement in childhood and taper sharply at eighteen; adult antiretroviral therapy services are configured for people who acquired HIV sexually and rarely accommodate youth treated since infancy.

A considerable number of youth with perinatally acquired HIV in India also spend part of childhood in residential childcare institutions (CCI) following parental death, family poverty, and stigma that makes kinship fostering unavailable.^4^ For them, discharge from CCI care at age 18, and transfer out of pediatric services arrive together, often without aftercare planning. ^5^ Existing transition readiness instruments were developed in high-income settings and operationalize readiness as individual autonomy focusing on medication self-management, independent clinic navigation, self-advocacy.^6^ Few were constructed with input from youth in low- and middle-income country (LMIC) settings, and few capture institutional transfer processes, family involvement, and community stigma.^6,7^ This study examines what readiness looks like when youth themselves define it.

Photovoice offers one route to that question. Introduced by Wang and Burris, it is a community-based participatory method in which participants photograph their surroundings, and collectively analyze the images, generating evidence about structural conditions shaping health.^8,9^ While its suitability is documented in populations where stigma constrains verbal disclosure, its use among YLHIV in South Asia remains sparsely documented.^10^ At the outset, a participant in Bengaluru asked: “what is this, and why are we doing it?” This paper is the extended answer: what young people said readiness requires, and what peer-facilitated photovoice made available.

## METHODS

### Context

The study was part of a mixed-methods multi-institutional initiative to develop a community-informed transition readiness assessment tool for YLHIV in LMIC settings. The photovoice component was conducted in March 2026 at two sites in Karnataka: Bengaluru, the state capital, and Belgaum (Belagavi), a mid-sized city in northern Karnataka. Sites were selected to capture variation in urbanity, health infrastructure, and living arrangement. Eligibility included YLHIV who had transitioned or were preparing to transition from pediatric to adult care. Sixteen participants were enrolled through purposive sampling from clinics and known networks (Bengaluru, n=8; Adults (18-22y), family-based or with prior or current CCI experience; Belgaum, n=8; Minors (13-17y), all residing in CCI). Trained peer facilitators included young adults with lived experience of HIV and CCI care who contributed to study design, implementation, and data interpretation.

#### Photography Training and Peer Facilitation

Participants attended a structured one-hour orientation co-facilitated by the research team and peer facilitators. Training used example photographs from other photovoice studies involving people living with HIV followed by interactive discussion and a caption-writing exercise. Peer facilitators modeled ethical practice including consent, privacy, and prioritizing safety over taking pictures.^11^ Participants were asked to take 1-3 photographs, each with a caption, reflecting their experience of growing into adulthood and their thoughts about the future. They were given one week between orientation and discussion to carry this question into their own lives, and to observe, photograph, caption, and reflect on their own terms. Focus group discussions (FGD) were conducted at each site. Each participant presented their photographs and their reflections guided by the SHOWeD (See, Happening, Our lives, Why, Do) sequence, positioning participants as interpreters of their own images rather than respondents to researcher prompts.^8^ The Bengaluru FGD was co-led by peer-facilitators, and conducted in English and Kannada with real-time translation. In Belgaum, peer facilitators assumed the primary facilitation role, with the researcher providing supportive oversight.

### Consent

Participants ≥18 years provided written informed consent; minors provided child assent following written parental/guardian permission. All documents were available in English and Kannada and administered in participants’ preferred language. Separate written consent will be obtained before any public exhibition, allowing participants to choose their attribution and images.

#### Member Checking

As part of data validation, following theme consolidation by the research team through the analysis described below, six participants from both sites joined a member-checking session by video conference, facilitated bilingually in English and Kannada by peer facilitators. Themes were presented for participants to confirm, correct, elaborate, or dispute ^12–14^. Reflections from this session are attributed accordingly in the Results.

#### Reflective FGD

In May 2026, after the primary photovoice FGD, the researchers conducted a reflective FGD with six participants (three per site) to elicit participants’ interpretations of the photovoice process itself. They were invited to reflect on how the presence and roles of researchers and peer facilitators may have shaped what they felt comfortable expressing.

#### Analysis

Analysis proceeded in two stages: coding of focus group transcripts and captions, followed by participant validation of the resulting themes. Translation and coding decisions were made collaboratively by researchers and peer facilitators. FGD transcripts were translated from Kannada to English by a bilingual researcher. Transcripts and photograph captions, including those from the member-checking and reflective sessions, were coded in Dedoose (v9) using a hybrid deductive-inductive approach.^15,16^ Coding domains were derived deductively from the FGD guide and transition literature, additional codes were generated inductively from participants’ photographs and discussions, allowing unanticipated concepts to emerge. Two researchers independently coded all transcripts. Inter-coder reliability was assessed in Dedoose.^15^ Pooled agreements were iteratively assessed, coding rules were clarified and transcripts recoded; final pooled Cohen’s kappa was 0.66 across 62 excerpts, indicating substantial agreement.^17^ Remaining disagreements were resolved by consensus.

## RESULTS

Primary analysis yielded four interrelated themes describing how participants understood growing up with HIV, what made transition feel possible, and what they needed to reach independent adulthood. A separate analysis of the reflective FGD examined their interpretations of the photovoice process. Illustrative quotes for each theme appear in Table 1.

### Independence as Relational Responsibility

Participants described independence as deeply relational. Becoming an adult was not primarily about managing one’s own medication, making decisions alone, or living apart from caregivers. It was about being able to support parents, repay caregivers, care for siblings, and contribute to the community’s wellbeing. Education, employment, and financial stability were framed as pathways to giving back rather than as personal achievements.

This obligation was articulated most fully through photographs. One participant photographed a tree, framing parental care as a debt discharged through how one lives (Figure 1). Worth was also socially conferred: another used the image of a flower to convey that a life has value when its growth is recognized by others. The relational orientation extended even to safety: one participant described wearing a helmet not only as self-protection but as protection of his parents from the grief of losing a child.

**Figure 1.**
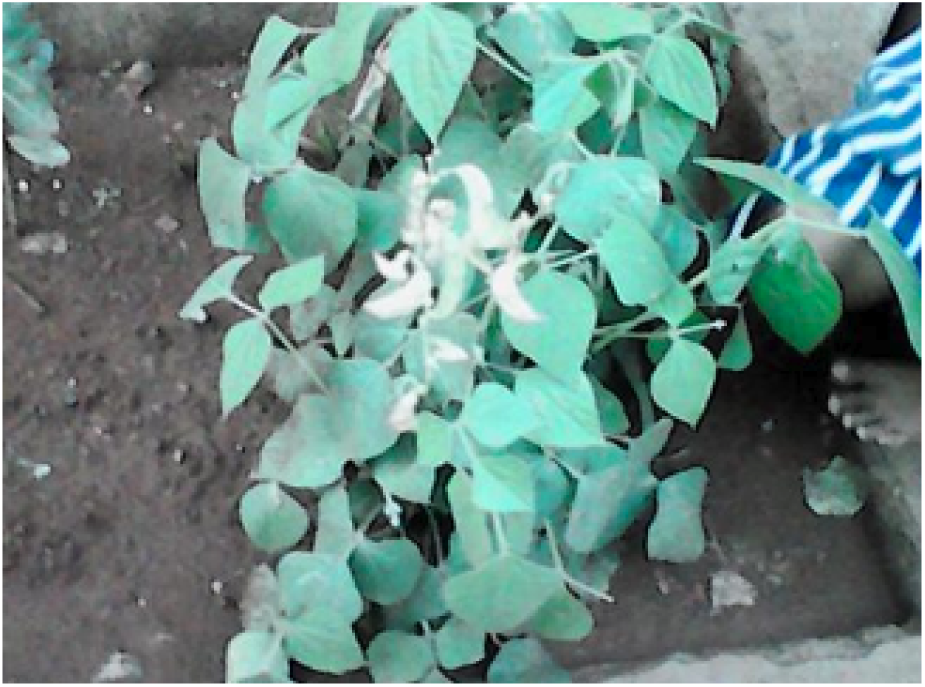
Tree. “A human should learn by looking at trees, because to grow a tree, you need water, fertilizer, and other inputs. Similarly, to raise a child, a mother has to work hard. After that child grows up, they should take care of the nation’s progress. Looking at the tree, that child should stand on their own feet and lead their own life.” - Belgaum participant, Photovoice caption, Female, Minor (< 18 years)

### Transition as Multidimensional Readiness

Consistent with this relational framing, participants understood readiness as extending well beyond HIV knowledge and adherence. Clinical self-management mattered, but education, housing, psychological support, and social preparedness were described as equally critical and largely neglected. Employment was a route to both self-sufficiency and reciprocity toward family.

Emotional preparedness emerged as central, and participants named it *peace*: the ability to slow down, make sense of one’s emotions, and stay balanced within circumstances one could not control. One participant photographed plants growing in constrained ground to convey this (Figure 2). Participants identified age 18 as a turning point, particularly for those leaving institutional care, but argued that preparation should begin around age twelve, framing transition as a gradual accrual of responsibility rather than a single event.

**Figure 2.**
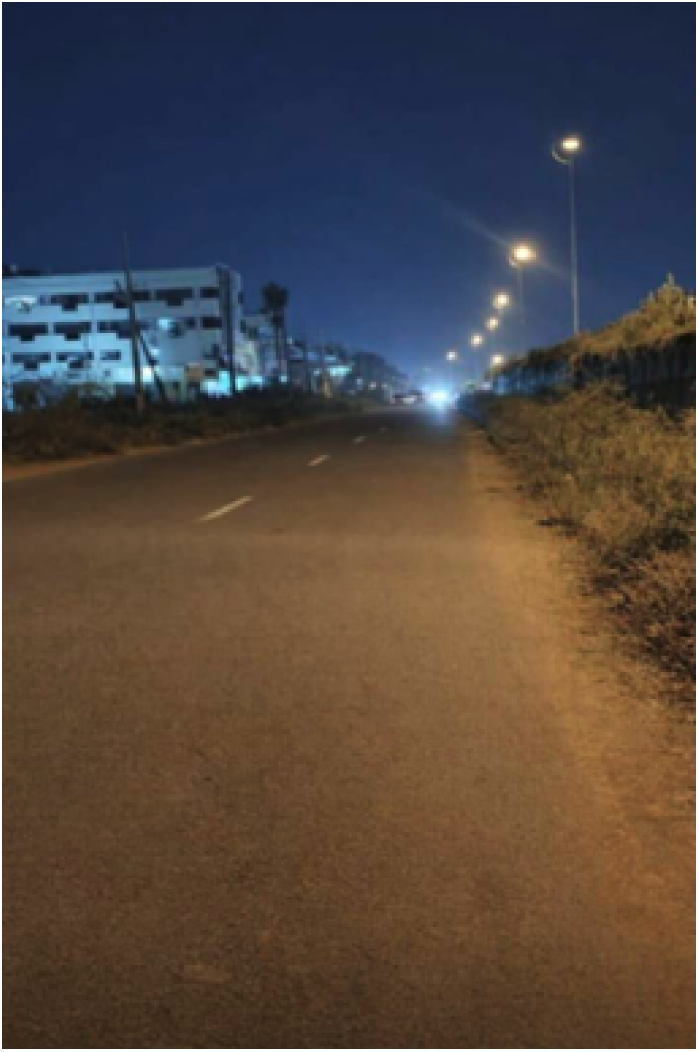
Road at Night. “In this picture, I have captured a beautiful scene from nature. When I saw this view, I felt very happy. It reminded me of how peaceful and meaningful nature can be.” - Bengaluru participant, Photovoice caption, Female, Adult (≥ 18 years)

### Anticipatory Loss of Home and Trusted Care

Transition was anticipated as loss before it was anticipated as independence. Family members, institutional caregivers, counselors, providers, and peers were described as a care network attuned to HIV-specific needs. In Belgaum, one caregiver, “*Madam*,” was at once a disciplinarian, confidante, and proxy parent, and her anticipated loss at discharge was a source of pronounced grief. The repeated sentiment that “no one will” care for them long-term reflected anxiety not of solitude but of unsupervised risk. One participant captured this through the metaphor of a sheep without a shepherd, framing caregiver support as protective and life-sustaining (Figure 3).

**Figure 3.**
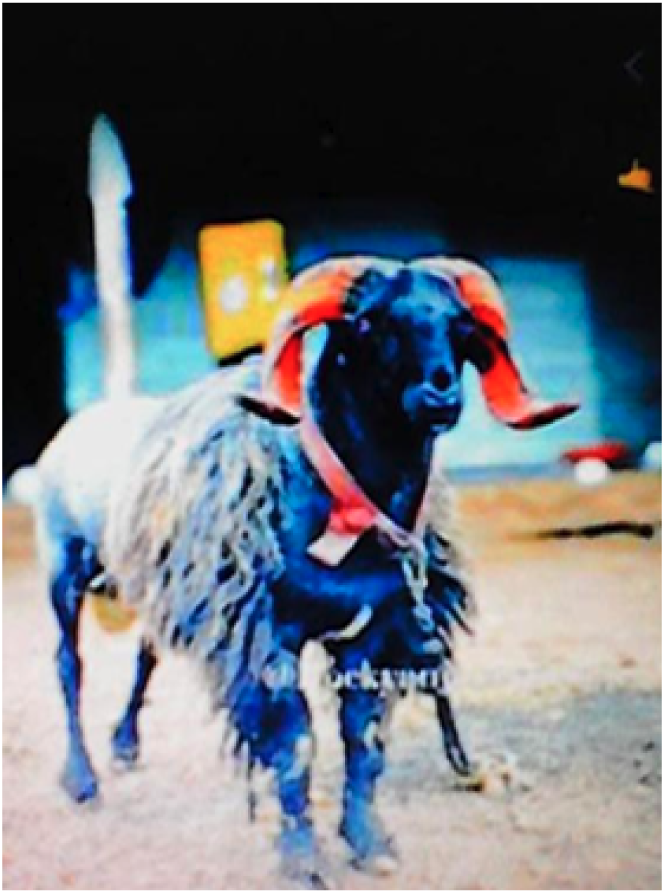
Sheep. “A sheep without a shepherd is lost and will go anywhere and do anything, and will put its life in danger. So, how a sheep needs a shepherd, we need a shepherd in our life. Otherwise, like the sheep, we will go anywhere and get spoilt, and we might lose our life.” - Belgaum participant, Photovoice caption, Male, Minor (≥ 18 years)

Participants who had already transitioned described these networks as attenuated rather than severed, continuing to draw on family and community for emotional and HIV-related support, suggesting the value of allowing supportive relationships to persist informally after transition.

#### Stigma and the Social Conditions of Transition

Stigma constrained help-seeking even where support existed. Fear of unintended disclosure limited what participants would raise even with trusted figures and produced a persistent awareness that their path to adulthood was shaped by HIV. Coping strategies included non-disclosure, cognitive reframing, spiritual coping, and stoicism; these appeared to hold down distress rather than resolve it. Others described a future-oriented coping, treating social judgment as an obstacle to be navigated rather than derailment of their aspirations. One participant depicted this through an image of steps (Figure 4).

**Figure 4.**
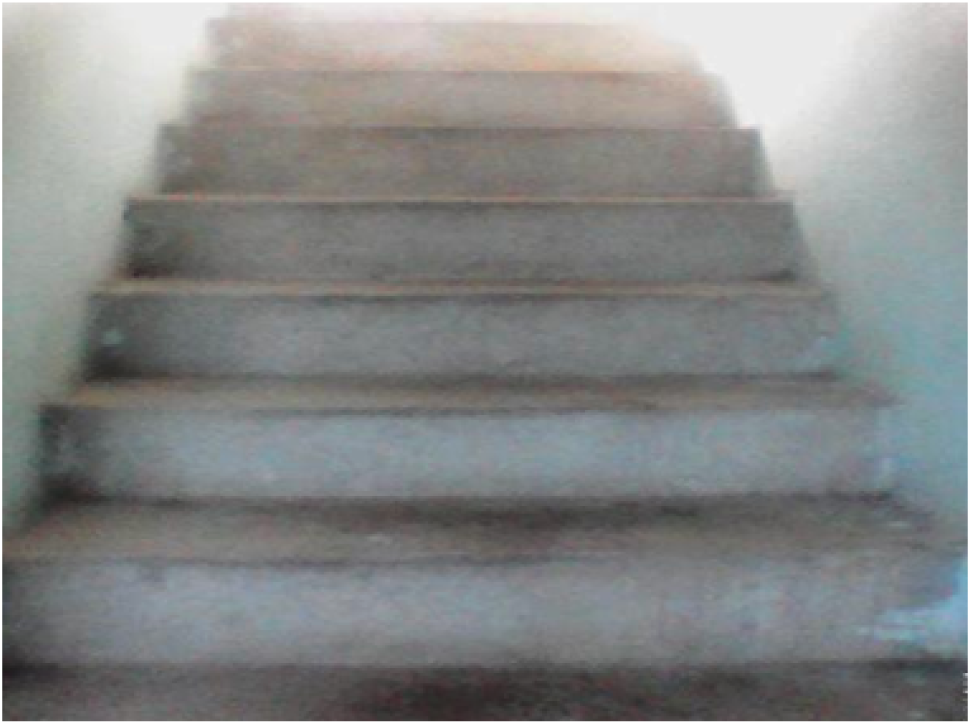
Steps. “I will attain my goals no matter the obstacles that come in my way. No matter what people say about me, I believe I will attain my goals. I will reach my aims.” - Belgaum participant, Photovoice caption, Male, Minor(≥ 18 years)

Participants did not accept stigma as an individual burden to be managed. They identified community-facing education, directed at adults rather than at youth, as the change required to make disclosure safe.

#### Participants’ Interpretations of the Photovoice Method

Participants described photovoice as helping them recognize and articulate experiences that had remained unseen. One participant reflected on a roadside market, where the unnoticed stories of everyday life became a way of thinking about marginalization more broadly (Figure 5). For YLHIV whose own experiences and efforts had frequently gone unrecognized, the photograph offered a symbolic language through which invisibility could be expressed.

**Figure 5.**
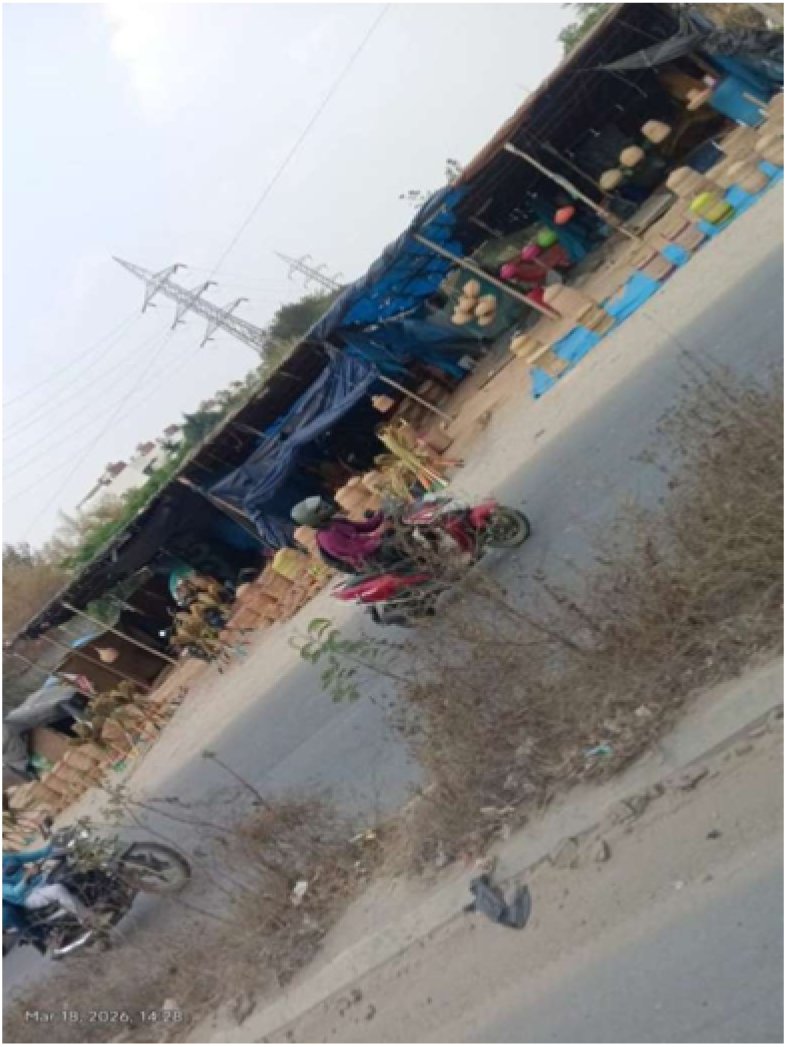
Roadside Market. “This photo shows a roadside market where handmade baskets and goods are sold, while life moves around it. It matters because it reflects the quiet hard work behind livelihoods that go unnoticed. This connects to life because it reminds me to value the effort, culture, and stories behind ordinary places.” - Bengaluru participant, Photovoice caption, Female, Adult (≥ 18 years)

Participants assessed photovoice more broadly as a powerful form of storytelling capable of making marginalized experiences visible.

*“….. I feel photo is a very strong tool for storytelling that would help to raise awareness and highlighting our personal experience and resilience of people among the vulnerable community that would help to reduce stigma.”* -Bengaluru participant, Reflective FGD, Male, Adult (≥ 18 years)

When asked about the power dynamics of whether the presence of senior researchers or older peers had affected their willingness to speak, responses varied. One participant rejected the premise, describing a conversation that gave them “freedom to express [their] ideas” (Table 1). Others were less categorical, but none described discomfort. With regard to older peers, participants attributed this comfort to facilitators’ familiarity with their language and daily lives.

## DISSEMINATION

### Returning the Images to the Community

Participants endorsed public display of their photographs. The images will be exhibited at a youth conference in late 2026, to which district health officers, program managers, clinical and institutional staff, and community members will be invited, returning participants’ analysis to the actors positioned to act on it.

## REFLEXIVITY AND POSITIONALITY

### Partnership Structure and Authority

This study was conducted with two South India-based organizations with long-standing community relationships: YRGCARE, which coordinated local research oversight, and RISHI Foundation, which supported peer facilitators and held the relationships with study sites. Both are represented in the authorship of this paper, as are the lead youth peer facilitators, themselves YLHIV.

### Peer Facilitator Positionality

Peer facilitators (sharing with participants a diagnosis, a language, and an experience of institutional life) completed written reflections before and after facilitation on how their identity might shape their interactions with participants and their interpretation of participants’ accounts. Their reflections identified two risks. The first was over-identification, the assumption that shared experience meant shared meaning:

*“Even when I think I understand what participants are trying to say, I should give them the opportunity to explain it themselves and confirm my understanding. It is important for me to represent their voices accurately rather than adding my own interpretation to their experiences*.”

- Peer Facilitator, Written reflection, Female, Adult (≥ 18 years)

The second risk was that peer status relocates power rather than eliminating it:

*“Even if I see myself as a peer or community member, I still hold power because I am asking the questions, recording information, and representing the research team. Participants may believe that my opinions or decisions could influence future services or support, which may affect what they choose to share.”*-Peer facilitator, Written reflection, Male, Adult (≥ 18 years)

### Academic Team Positionality

This study addressed questions youth had already named as urgent; the academic contribution was methodological rather than agenda-setting. The academic authors are researchers based in the United States and India, working in HIV and implementation science, none with lived experience of growing up with HIV or of institutional care. Several were familiar with existing transition readiness frameworks, an exposure that predisposed us to look for the individual-autonomy constructs those instruments measure, and made participants’ relational account initially appear anomalous rather than definitional. This shaped the deductive portion of the codebook, and the study’s central finding required revising an assumption we had not recognized as one.

## DISCUSSION

The central finding was that participants defined transition readiness and independence as the capacity to sustain others rather than individual autonomy, a dimension largely absent from existing instruments. Readiness tools developed in high-income settings operationalize preparedness as individual autonomy: independent medication management, unaccompanied clinic navigation, and self-advocacy.^6^ Participants instead described reciprocal obligation: the capacity to give back, to repay parents, provide for siblings, and become a source of support within the networks that had raised them.

This relational construct organized every other dimension of readiness. Clinical self-management mattered, but as one strand among educational, employment, psychological, housing, and social preparedness. Employment was valued because a salary supports a sibling or other family members; emotional steadiness, which participants named as *peace*, was valued because instability jeopardizes the capacity to be relied upon. Participants framed transition as a gradual accrual of responsibility rather than an event tied to a clinical or institutional threshold.^18,19^ They proposed that preparation begin at approximately age twelve converging with international guidance recommending initiation in early adolescence despite reporting no structured preparation at any age.^18^

Participants located the principal threats to transition outside the clinical domain. Those approaching discharge described leaving CCI care as a loss rather than as a gain in independence, with grief centered on the caregiver who had been most consistently present. Stigma constrained what they could discuss even with supportive family and friends, prompting coping strategies such as non-disclosure and stoicism, that contained distress rather than resolving it.

### Who Facilitates Shapes What Is Learned

Participatory research redistributes interpretive authority rather than merely soliciting participant input, consistent with Freirean critical consciousness and with established CBPR practice.^8,20–22^ Fournier and colleagues found that peer co-facilitation with children living with HIV in Uganda was essential to ethical integrity and participant trust,^23^ and existing photovoice literature has similarly emphasized the ethical and relational dimensions of peer involvement.^11,24^ Our findings suggest it also affects epistemic yield: what participants say, and therefore what a study is able to find.^18^

Three observations support this. Reciprocal obligation was absent from the initial codebook and surfaced through participants’ repeated visual metaphors; this was a construct the academic team’s prior exposure to autonomy-based frameworks had not anticipated. Peer facilitators’ knowledge of the vocabulary, social hierarchies, and porous boundary between belonging and surveillance within residential care allowed facilitation to be calibrated in ways an outsider team could not. And participants attributed their own comfort to facilitators’ familiarity with their language and daily lives, describing the sessions as informal and conversational.

The images and captions producing the relational-independence finding came mainly from Belgaum, where peer facilitators led and the academic team stepped back. The sites differed in participant age, residential setting, and session format, so no controlled comparison can be drawn; we offer this as a hypothesis for studies designed to test it.

One younger participant offered a necessary qualification, describing the photography as difficult at first and rewarding only afterward. Photovoice is not equally easy for everyone: composing an image and explaining what it means requires visual and narrative confidence that younger adolescents may need facilitation to build rather than assume.

### What Peer Facilitation Requires

Peer facilitation should be treated as a methodological requirement rather than an optional enhancement. Peer facilitation can foster participation and collective meaning-making, while facilitators’ lived and contextual knowledge can strengthen culturally grounded engagement and interpretation.^22,25^ Over several days our facilitators listened to accounts of stigma, caregiver loss, and institutional discharge closely resembling their own histories. Facilitating that material is emotionally demanding and should be resourced accordingly, through supervision during the study, structured debriefing afterward, and payment commensurate with the role.

It also depends on relationships that predate the study. Peer facilitators cannot be recruited at the protocol stage. They must already be known to an organization that has supported them long enough to identify who is willing and prepared, and that can continue supporting them once the research ends. Studies without such a partnership should be cautious about claiming the participatory standing that peer facilitation confers.

Peer status also does not automatically resolve the power asymmetry it is invoked to address. Our facilitators recognized that their perceived proximity to a service-providing organization could constrain participant candor, suggesting that peer facilitation may not completely dissolve power. The appropriate inference is not that the model is compromised but that it must be documented and interrogated rather than asserted as a participatory credential.

### Limitations

The small sample size (n=16 across two sites in one Indian state) may have limited transferability. The reflective FGD for photovoice assessment involved a subsample (six of 16 enrolled participants). Site-level differences in participant age and residential status may confound the facilitation comparison described. Member checking was conducted online, and for several Belgaum participants this was a first videoconference, introducing a mode effect that in-person verification would not carry. Belgaum participants were recruited through the institution in which they resided, and we cannot exclude the possibility that this dependency shaped what was said about institutional care.

## PUBLIC HEALTH IMPLICATIONS

### Measure transition readiness broadly and early

Transition readiness measures for LMIC settings should assess reciprocal obligation to family, anticipated loss of trusted caregivers, and disclosure safety, alongside clinical self-management, and should treat educational, employment, and housing preparedness as constituents of readiness rather than context. Instruments measuring clinical competence alone will classify as ready those young people still facing the risks identified here. Programs generally taper psychosocial support at a chronological threshold, with limited adolescent-specific provision.^26^

### Stigma as a socially addressable target

Participants framed stigma as a community problem, not an individual burden. Current programs address stigma primarily through individual coping support, which remains necessary but limits change.^27,28^ Programs should pair individual support with community-facing education directed at families, employers, school staff, and health workers outside HIV services.^29^

### Refine reporting standards for participatory research

Participatory and extractive photovoice are procedurally indistinguishable as both involve cameras, captions, and group discussion. What separates them is who interpreted the images. Journals should require reports to state: (1) who facilitated image discussion and their relationship to participants; (2) who generated the initial codebook; (3) whether participants reviewed or revised the themes; and (4) whether participants’ input changed any analytic conclusion, and at what stage.

## CONCLUSION

Participants articulated a definition of adulthood that existing readiness instruments do not accommodate: becoming independent meant becoming able to sustain others, and the risks of transition were relational and social before they were clinical. Photovoice made that definition available in ways conventional qualitative inquiry would not. It relocated the research encounter into participants’ own surroundings, supplied a medium through which stigmatized experience could be shown before it was spoken, and positioned participants as analysts of their own images. Peer facilitation of the photovoice method was a pre-condition that made this possible. These methodological gains generated practical recommendations: participants named stigma as a community problem requiring community response, specified when transition preparation should begin, and identified what a readiness instrument must measure for accuracy.

Photovoice generated evidence that participants authored, can stand behind, and can use to advance their own case for change. That is the answer to the first participant’s query: this is what it was, and why we were doing it.

## Supporting information

Table 1. Illustrative Participant Quotes by Theme

## Data Availability

All data produced in the present study are available upon reasonable request to the authors

## ABBREVIATIONS

AIDS: Acquired immunodeficiency syndrome
ART: Antiretroviral therapy
CBPR: Community-based participatory research
FGD: Focus group discussion
HIV: Human immunodeficiency virus
ICR: Inter-coder reliability
IRB: Institutional review board
LMIC: Low- and middle-income country
SHOWeD: See, Happening, Our lives, Why, Do
YLHIV: Youth living with HIV

