## Supplementary material for "Peer Facilitation as a Methodological Condition: What Participatory Photovoice Revealed About Transition to Adulthood Among Youth Living with HIV in India": Table 1. Illustrative Participant Quotes by Theme

| Theme | Illustrative Quote | Source |
|---|---|---|
| **Independence as Relational Responsibility** | *"From the day we were born, parents have let us be free. Until we turn 18, we live freely…We should work hard, earn, do good for our family, give them freedom too, and give them the satisfaction of having raised us."* | Belgaum participant, FGD, Female, Minor (<18 years) |
| | *"Our parents grow us like a flower & show us the way to work. We have to live by their word. In case the flower dies, we also lose our life."* | Belgaum participant, Photovoice caption, Male, Minor(<18 years) |
| | *"When riding a vehicle, we might accidentally fall, and our head could get hurt, the brain could be damaged, and we could be in a situation where our parents lose us..."* | Belgaum participant, FGD, Female, Minor (<18 years) |
| **Transition as Multidimensional Readiness** | *"I like to go to a job which gives me a high salary to handle my daily needs to support myself and my brother also, because he is supporting me now so, I have to get good job."* | Bengaluru participant, FGD, Female, Adult (≥ 18 years) |
| | *"From 12 years itself, we should start taking responsibilities. By the time we're 18, responsibilities will have increased and we should continue carrying that responsibility."* | Belgaum participant, FGD, Female, Minor (<18 years) |
| **Anticipatory Loss of Home and Trusted Care** | *"Leaving the hostel is kind of happy and kind of sad. Especially leaving Madam, that will be truly sorrowful."* | Belgaum participant, FGD, Male, Minor (<18 years) |

| | | |
|---|---|---|
| | *"My mother and [institution] family, whenever I am lonely and distressed my mother is always there to help me... when I was afraid of HIV and all and the things running in my head."* | Bengaluru participant, FGD, Female, Adult (≥ 18 years) |
| **Stigma and the Social Conditions of Transition** | *"In the later stage no one will care about you because after your parents, no one will support you... Sometimes we can't even share it with our friends because we can't talk to them about our medicines and other things."* | Bengaluru participant, FGD, Male, Adult (≥ 18 years) |
| | *"The difficulties that come as we grow; we shouldn't share them with anyone. We should swallow our difficulties and keep going."* | Belgaum participant, FGD, Female, Minor (<18 years) |
| | *"This transition is not only needed for youth... it all depends upon the social attitudes... things like stigma... So, there should be certain education or awareness created... Not just for young people, but also for adults and old people."* | Bengaluru participant, FGD, Male, Adult (≥ 18 years) |
| **Participants' Impressions of the Photovoice Method** | *"Taking the photos was a bit difficult at first, but the experience was good and it made me happy."* | Belgaum participant, Reflective FGD, Male, Minor (<18 years) |
| | *"No, I don't think age is the problem. Because we were totally comfortable in the photovoice session-it was not a formal conversation, it was completely in a friendly manner. So, I think we had freedom to express our ideas."* | Bengaluru participant, Reflective FGD, Male, Adult (≥ 18 years) |

| | | |
|---|---|---|
| | *"I think all the things we said will be very useful for others."* | Belgaum participant, Reflective FGD, Male, Minor (<18 years) |
